# Medical Students’ Perceptions of Learning and Working on the COVID-19 Frontlines: “… a confirmation that I am in the right place professionally.”

**DOI:** 10.1101/2021.12.01.21267145

**Authors:** Jennifer M. Klasen, Zoe Schoenbaechler, Bryce Bogie, Andrea Meienberg, Christian Nickel, Roland Bingisser, Kori LaDonna

## Abstract

**Introduction:** The COVID-19 pandemic caused complex and enduring challenges for health care providers and medical educators and changed the medical education landscape for learners. Medical students were required to adapt and learn in a novel learning environment while universities paused their formal medical training. The current study sought to investigate medical students’ experiences working on a pandemic frontline to understand how they perceived this novel learning environment influenced both their learning and their developing professional identity.

**Methods:** We conducted semi-structured interviews with 21 medical students who worked in a COVID-19 testing facility at the University Hospital of Basel. Using constructivist grounded theory methodology, we collected and analyzed data iteratively using a constant comparative approach to develop codes and theoretical categories.

**Results:** Participants described improvements in their technical and communication skills, consequently impacting their professional development. The presence of a perceived flat hierarchy between the physicians and medical students promoted professional identity development amongst the medical students. Most participants perceived working on the pandemic frontlines as a positive learning experience, which seemed supported by a flatter hierarchy and open communication compared to their usual learning environment.

**Conclusion:** Since medical students reported that their work on the pandemic frontlines positively affected their learning, the need to create hands-on learning opportunities for medical students challenge curriculum developers. Medical students wish to feel like full-fledged care team members rather than observing learners. Performing simple clinical tasks and collaborative moments in a supportive learning environment may promote learning and professional development.

## Introduction

Not surprisingly, many countries postponed all in-person medical education in response to the COVID-19 pandemic [1] [2]. Consequently, traditional medical education and training were either cancelled or shifted to virtual platforms, causing many medical students to miss out on formative bedside teaching opportunities [3]. This drastic perturbation in normal day-to-day operations not only created uncertainty for medical students, but also fostered persistent concerns that their learning and professional development would suffer [4]) [5] [6]. Although medical students have historically been sidelined during public health crises such as the 2003 SARS epidemic [7] to prevent disease transmission and to relieve staff from their teaching commitments [8], the role of medical students in this pandemic remained unclear and authorities handled it differently [9]. Medical students in Basel, Switzerland were invited to serve as first responders on the COVID-19 frontlines [10], much like during the Spanish Flu in 1918 or in the current pandemic in Vietnam [11] [12]. Our goal was to understand how medical students’ experiences in this novel clinical learning environment might inform post-pandemic medical education.

The Corona Task Force (CTF) of the University Hospital of Basel (UHB) created a Triage Test Centre (TTC) to relieve some of the healthcare system pressures caused by a high flow of incoming patients and a corresponding shortage of personnel and resources [13]. Medical students were recruited to join a specialized response team called the “SWAB team” [10] where, with appropriate safety protocols in place, they supported senior health care workers by assisting in the clinical evaluations of hundreds of patients per day. These evaluations included taking medical histories, performing nasopharyngeal and oropharyngeal swabs, and providing counselling to patients about COVID-19 behavioural guidelines [10].

The TTC afforded medical students an experience vastly different from traditional curricula. In Basel, the traditional curriculum integrates some hands-on practices from the curriculum’s beginning (Fig 1).

**Figure 1:**
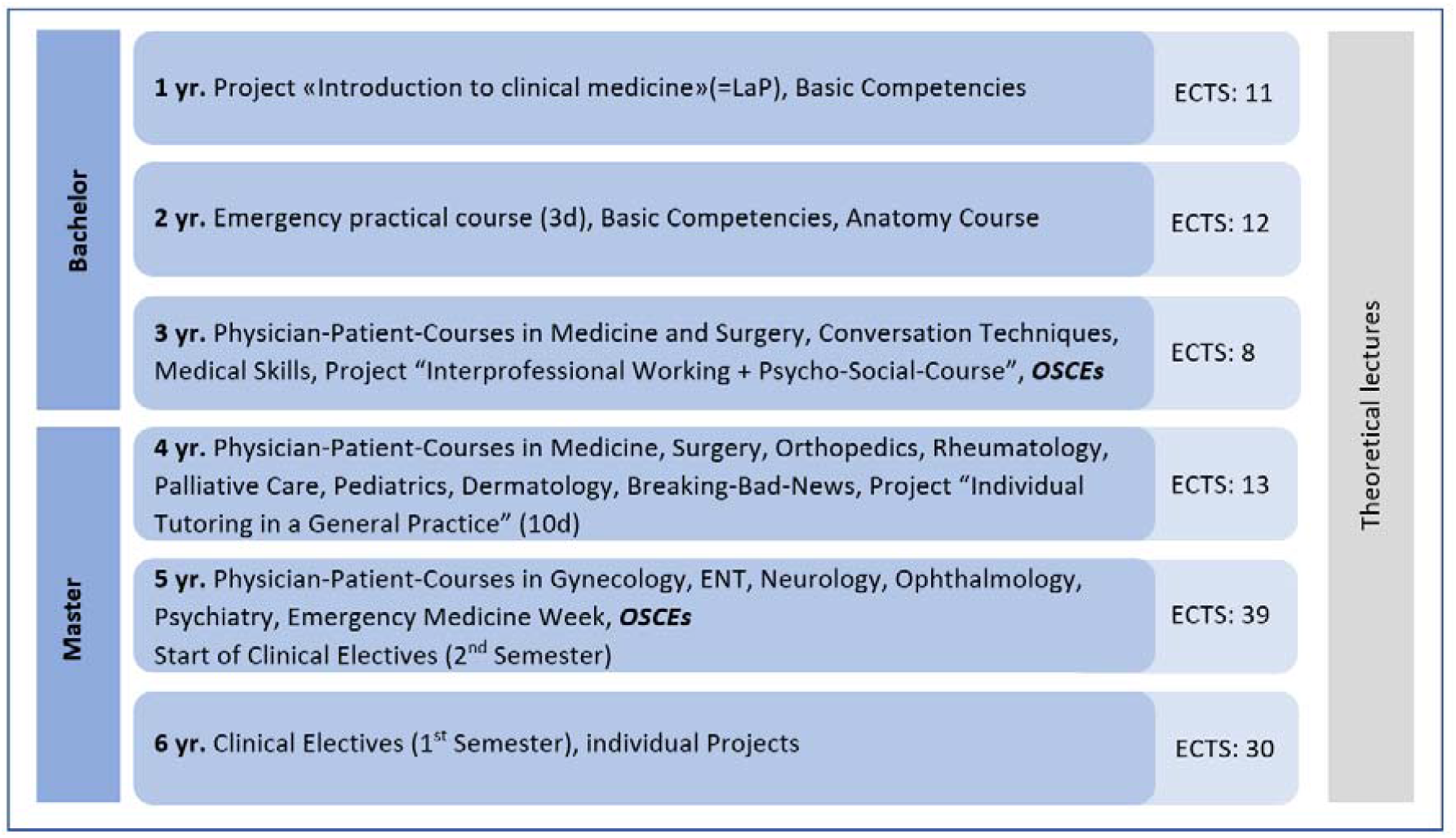
Clinical and Hands-on Courses at the University of Basel. (Detailed description: Medical students gain their first clinical experiences through a format called Learning on the project (LaP) in the first year. LaP includes several bedside-teaching sessions and other learning opportunities in the hospital such as observing surgery. Before or during the first year, the University also expects a month-long clerkship as a nurse assistant in a hospital. In the second year, medical students work three days in the ED, guided by nurses. From the third year to the fifth year, Basel’s medical students participate in the “physician-patient-courses” to learn conducting a medical history and physical exams. Those small group learning sessions cover some of the most critical fields in medicine, closely supervised. In the fourth year, medical students observe a family doctor to collect further professional insights. From the fifth to sixth year, medical students work as full-time clerks in the hospital for one year. As the final exams of the third and fifth year, *objective structured clinical examinations (*OSCEs) are conducted to monitor their clinical competence skills and development.)

However, in medical schools around the world, medical students—particularly those in their first few years—have limited, hands-on interactions with patients, and they rarely play a collaborative role on the care team [14] [15]. Although observation is a useful pedagogical activity for medical students [16] [17], opportunities such as medical student-run health clinics with numerous patient interactions provide an invaluable educational opportunity [18] [19].

Physicians—particularly those tasked with preparing trainees to become competent providers—are constantly striving to improve the quality of education and care. Since extraordinary circumstances might reveal insights useful for transforming teaching and practice, we explored medical students’ experiences working in a learning environment necessitated by a once-in-a-generation crisis. By examining their perspectives, we may identify the features of this novel setting that medical students perceived as particularly influential for skill acquisition and professional development.

## Methods

We conducted semi-structured interviews with medical students who volunteered in the TTC during the first wave of the COVID-19 pandemic to explore their perceptions of learning and working on the frontlines [20]. This study adhered to the Declaration of Helsinki and the Ethics Commission Northwest and Central Switzerland (EKNZ, Req-2021-00518).

### Study context

The Preachers ‘ Church, where the TTC was located, operated from March to June 2020. The TTC was staffed with physicians, nurses, other healthcare providers, army staff, and medical students who worked on the “SWAB team”. Two shifts were scheduled per day consisting of 12 medical students each. This schedule translated to a total of 936 shifts that were covered by medical students in March and April 2020. All medical students received a paid working agreement and appropriate insurance coverage. The SWAB team curriculum included: history taking; testing with swabs; learning about social distancing and disinfection; and hygiene, self□protection, and careful handling and donning of PPE which was provided by experienced physicians who acted as clinical supervisors on-site. There were no formal learning objectives associated with this clinical placement, nor were there any formal examinations required of the medical students.

### Sample

We purposively sampled participants through both word-of-mouth communication and through the TTC chat group. Because they’d worked at the TTC from its inception and had previous bedside training experiences, we initially sampled medical students in years 4-6 of the traditional Basel curriculum. These participants recommended other possible interview candidates, so we modified our recruitment efforts to include snowball sampling. As the iterative data generation and analysis progress unfolded, we also theoretically sampled participants to ensure diversity in both gender and level of training. We recruited participants until we reached theoretical sufficiency.

### Study design

Participation was voluntary and we obtained written informed consent before each interview. Interviews began with an easing-in strategy, asking participants about their previous medical experiences and their motivations for working on the COVID-19 frontline. As the study progressed, the interview guide was modified to include questions about specific learning experiences at the TTC, as well as questions pertaining to professional identity formation. ZS conducted the interviews from May to October 2020. The interviews were held via Zoom (*n* = 17), Skype (*n* = 1), telephone (*n* = 2), and in-person (*n* = 1) using PPE and in accordance with public health guidelines. The interviews lasted between 30 and 70 minutes.

### Data analysis

All interviews were audio-recorded in Swiss German, and then ZS transcribed and translated each recording to German. All identifying information was removed from the transcripts. Using a constant comparative approach, we identified and refined codes and categories by analyzing data both within and across transcripts [20] [21]. Specifically, ZS read all the transcripts, while JK read the first seven in full. During frequent meetings, ZS and JK discussed the data and developed initial, focused, and theoretical codes [20]. To support the evolving analysis, team members (JK, AM, BB, and KAL) reviewed rich transcripts and data excerpts selected by ZS; of note, ZS translated representative selections of findings into English for KAL and BB to review. Quirkos Version 2.3.1 was used to manage the data. ZS wrote field notes and analytical memos that were instrumental for informing the analysis.

### Research team

ZS is a medical student at the UHB who worked at the TTC. Influenced by her experiences, ZS conducted her master’s thesis on learning and working amidst the COVID-19 pandemic. ZS’ written reflections prior to the study supported reflexivity, helping her recognize personal beliefs and acknowledge potential influences. JK, the principal investigator of the study, is a senior surgeon at the UHB who is interested in medical education research. JK incorporated her experiences and reflections as both a former medical student, resident, and current qualitative, medical education researcher and clinician teacher to guide her contributions to this study. AM, CN, and RB were involved in study design, and manuscript preparation, drawing on their knowledge and involvement in the development and management of the TTC. BB is a Canadian MD/PhD student with experiences working as a medical student on the frontlines of the pandemic in Canada. He has a diverse background in medical education research. He and KAL—a PhD trained scientist with expertise in qualitative methodologies— provided an international perspective on the current research, regularly providing feedback as the analysis evolved and assisting in the writing and critical editing of the manuscript. The team acknowledges that the personal and professional disruptions they experienced because of the pandemic likely influenced their interpretations of the data.

## Results

Twelve female and nine male medical students between 20-26 years of age and representing various stages of training participated in the study. Of these, medical students in years 4-6 (n=12) worked for around three to four months in the Preacher’s Church TTC, while the students in years 1-3 (n=9) worked there for one to two months. The frequency of workdays varied between one to six days a week, although most students worked three days per week.

### Learning, adapting, and improving skills

The learning environment at the TTC did not look or feel like a typical clinical setting:

*“For me, the atmosphere and mood in Preacher’s Church were exceptional. It is wonderful to see the cathedral with the pipe organs upstairs and then the people below who look like Martians in their white space suits”* (P1). Working under strict hygiene provisions was also a novel feature that seemed exciting—even other-worldly—for some participants: “*I was strangely dressed like that because I had never worn such complete protective equipment before. Somehow you felt mega special”* (P5).

Medical students across all years of training had to quickly adapt, not only to these startling surroundings, but also to engaging autonomously in both simple and complex clinical tasks. Some tasks, such as “taking the swabs” (P1) were generally perceived as easy to learn yet educationally impactful. Additionally, because they were given the opportunity to practice concepts they’d only understood theoretically, even seemingly mundane tasks were perceived as extremely valuable. For example, although all medical students are taught the importance of hygiene early in their training, following proper hygiene techniques such as disinfecting hands and surfaces seemed to crystalize learning: *“It certainly made me think about hygiene, how to properly dress and undress, what steps and in such a way that it really makes sense”* (P4).

Not surprisingly, this type of learning was particularly valuable for medical student participants in years 1-4, who reported that the opportunity to engage in frequent hands-on patient care drastically improved their diagnostic and technical skills. For instance, medical students reported learning how to identify and consider differential diagnoses for COVID-19:*”… In this sense, tonsillitis, bronchitis, pneumonia, or pulmonary embolism. And yet, every now and then, a suspicion of angina pectoris”* (P21) From their patient encounters, others learned the importance of not presuming a diagnosis too early:

> *“Another crucial experience was when … one day someone with lung cancer and someone with temporal arteritis were discovered by students, and I found that very impressive. There is another view* … *if something strikes you as suspicious, you look it up again”* (P7).

For both, 5^th^ and 6^th^ year medical students, tasks such as taking nasal swabs quickly became rote. However, although they perceived that they did not learn much that was new, their overall experience at the TTC reinforced previous learning and permitted them ample opportunities to practice various skills (P6, P7, P9). Indeed, all participants appreciated that, during busy days at the TTC when there were many incoming patients to consult, they were afforded considerable time and autonomy to improve their history taking, diagnostic, and technical skillsets. For instance, one participant remarked that working in a high-acuity setting under extraordinary circumstances helped them learn *“… how to organize a history taking efficiently and in a structured manner. … in the end, it was* … *a matter of 5 minutes. And not necessarily worse, you just knew what the crucial questions were”* (P8).

Perceived improvements in clinical skills resulted not only from directly performing these tasks, but also from engaging in observational learning such as listening to physicians and more senior medical students interact with patients: *“What’s more* … *not active, but passive, if you listen to certain physicians doing triage, I find it incredibly instructive”* (P21). Another student shared that: *“… listening to others taking a medical history, those are things that I have to master at some point, and just as they approach the situation and talk to patients”* (P5).

Indeed, for most of the participants, the greatest perceived improvement was not in the realm of technical skills, but rather in the development of so-called ‘soft skills’ like patient centered communication: *“At times it was just the knowledge of human nature or dealing with patients. I think that was almost the biggest point from a medical point of view”* (P11). Every participant reported improvements on the level of communication skills and the ability to provide individual care for patients: *“Precisely this routine and the feeling of how to approach people and deal with them”* (P2). Another participant described: *“That you had to do something that was just not pleasant and then pack it in a communicative way so that it was still okay for the patient”* (P3). Some participants reported that these experiences boosted their confidence in managing difficult patients, while others reported that they developed better techniques for obtaining critical information faster while still making the patient feel understood—a skill they perceived as invaluable for their future clinical practice (P3, P21).

### Interactions and flattened hierarchy

Mostly, participants appreciated “… all the interaction with colleagues” (P4), noting that feeling like an integral part of the team was the most striking and valuable feature of this learning environment. For participants, performing an essential role alongside peers, physicians, nurses, and military service members—rather than observing from the wings— was both atypical and formative: “*we really saw in this church that it doesn’t matter whether you are a student, a resident or a consultant, everyone can contribute and then a team works”* (P15). For participants, this sense of comradery broke down the hierarchical barriers governing their typical interactions with faculty in traditional learning settings: *“Especially at the beginning in the Church it was mega cool how flat the hierarchy was, and somehow everyone communicated fully openly and yes* … *it was just an enjoyable working environment”* (P17). Participant 15 agreed: *“I had the feeling that the hierarchy in the Preacher’s Church was lifted”* (P15). Not only did participants report that *“It was cool to talk to the doctors”* (P13) and *“it was always easy to talk to all physicians and yes, it was fun too”* (P8), many participants appreciated that the physicians made purposeful efforts to interact with them to promote their training: *“I also think that the doctors did it very well and took enough time* … *they said: ‘Look, we have time, we don’t have to do* …*’ and then they asked about our medical knowledge, which was fun”* (P8). The TTC learning environment even emboldened some participants to speak up or pushback, with one participant finding it remarkable not only *“… that you dared to speak up and even criticize junior and senior consultants”* (P4) but also that:

> … *other physicians were always delighted to receive feedback, like: “Hey, something is not going well there at the moment, can’t you try to change something?”* … *And many were really grateful for it, and that was nice*, … *And that helped me a bit to notice that most of them are satisfied when you address the problem”(P4)*.

While all participants obtained valuable learning, not all perceived that this experience was necessarily worth it:

> *“I would say that working at Preacher’s Church has undoubtedly brought me something for my studies. Still, to be honest, it would have been better for me if the COVID-19 pandemic had not broken out and we had continued our studies regularly”* (P1).

### Development of professional identity

For most, however, their experiences at the TTC were hugely influential for both facilitating their learning and charting their future career paths. For instance, some described that their work on the COVID-19 frontlines provided a “sneak-peek” into emergency or disaster medicine, piquing their interest in pursuing these specialities down the road (P2, P13, P17):

> *“I found it very exciting, and I could imagine going in this direction at some point later, in emergency medicine or something else, where something is always going on. I could still imagine that, and I don’t know whether I could have said that beforehand”* (P17).

However, for most participants, their work on the frontlines validated their choice to become a doctor, noting that their work in the TTC provided “*such a confirmation that I am in the right place professionally”* (P21). Another participant confirmed: *“It rather encouraged me that I think I will have a cool job that will give me back what I am looking for”* (P6). Participant 16 encapsulated the experience that all participants conveyed in various ways— working on the COVID-19 frontlines was invaluable because, for the very first time, they felt like *“patients saw [them] as a real doctor”* (P16).

## Discussion

We sought to investigate how medical students perceived learning on the frontlines during the COVID-19 pandemic. All participants reported that their experiences at the TTC were positive and effective in promoting the development of knowledge and task-oriented and intrinsic skills such as communication. These experiences further promoted the professional development of the medical students during a time of uncertainty about their current and future medical education and clinical experiences.

As a novel learning environment, the features of the TTC that facilitated learning seem to center around the opportunity to repeatedly perform various medical tasks while serving as an integral member of the care team. The different and changing assignments of medical students to specific tasks enabled a long duration for learning acquisition, repetitive practice with feedback, and memory consolidation [22]. The literature supports that features of the TTC such as hands-on engagement, even for seemingly mundane tasks like nasal swabs and disinfecting surfaces, provide significantly superior medical education and patient outcomes [23]. The discrepancy of learning between medical students from the first years to the senior medical students was likely a product of the traditional curriculum structure where junior learners rarely experience opportunities to perform tasks directly and with autonomy, nor do they have the opportunity to routinely observe physicians and senior medical students perform duties in real clinical settings [24] [25] [26]. However, autonomy is an esteemed sociocultural value in medicine rarely afforded to medical students, particularly those early in training [27]. The TTC therefore allowed junior learners to gain early exposure to the clinical environment, translating into perceived improvements in clinical knowledge and skills.

The conditions that were most conducive to optimal professional identity formation therefore occurred when the medical students were actively engaged in hands-on care, and thus, perceived they were engaging as service-providers and *not* as learners [28] [29]. In other words, medical student participants felt, often for the first time, that they were providing an essential service—a rarity in the traditional curricula, particularly for junior medical students. Since learning and professional identity development depend on active engagement [30], we and others call for clinician teachers and curriculum developers to more meaningfully embed opportunities for medical students to interact with patients, to autonomously perform tasks appropriate for their level of training, and to take part in team meetings at all points during undergraduate medical training [31] [32]. As we know that such learning opportunities for medical students differ from programs, countries and even regions, they shouldn’t be extraordinary, they should be standard, while we acknowledge the design of a curriculum might take a lot of creativity on the part, but the effort may pay dividends.

Interacting meaningfully as part of a team was vital for professional identity formation [33]. Traditionally, medical students are presented with a stricter learning environment with a more explicit and fixed hierarchy than that of the TTC environment [34] [35] [36] [37]. During regular bedside teaching sessions, medical students are guided by supervisors and have a clear schedule and defined tasks to complete when interacting with patients. Participants reported that the traditional hierarchy in pre-pandemic clinical environments collapsed into a “flat hierarchy” wherein the medical students perceived that they were viewed more as colleagues rather than learners. Perhaps the widespread uncertainty about the novel coronavirus, along with the common purpose of combatting COVID-19, fostered a shared sense of learning and adaptation amongst faculty and medical students. Even if the hierarchy wasn’t as flat as they perceived, perceptions matter. Feeling like a legitimate team member both fostered confidence and reinforced participants’ sense that they’d pursued the right professional path. In turn, medical students interacted with physicians and other medical staff differently than they usually would in the hospital setting, adopting a more casual style, and in some cases, pushing back and offering critique.

Another explanation could be that this sense of identity transformation from learner to healthcare provider is similar to the natural identity transformation that occurs when a medical student transitions from pre-clinical to clinical training [38] [39]. At the TTC, this transition happened much faster for junior medical students than the traditional curricula usually afford. Such a transition is indeed associated with increases in clinical responsibilities as medical students become more widely integrated into the clinical setting and care team. In the traditional curriculum, this transition is likely not associated with a complete flattening of the hierarchy, but more of a reduction in the natural hierarchy given the continued presence of fixed roles between medical teachers and learners. Under the unprecedented circumstances of a pandemic, learners had the opportunity to feel a sense of being a valued member of the team—providing a crucial service during a time of considerable uncertainty, which might influence the phenomenon of identity transformation from learner to healthcare professional.

Overall, through its design, the TTC accomplished these desired outcomes, establishing that, if such in-person, interactive learning is achievable amidst a catastrophic healthcare crisis, it could feasibly become part of the ‘new normal’ of post-pandemic medical education. While most medical students around the globe continued their training from behind a computer screen, the learners at the TTC were fostering their clinical prowess in an in-person environment that allowed them to *feel* like legitimate healthcare contributors.

### Limitations

This study reports the perspectives of medical students who consented to be interviewed about their experiences working in a novel learning environment. Findings may not necessarily reflect the perspectives of all the medical students who worked at the TTC. As a medical student herself, ZS conducted all interviews to engage her peers in a casual but candid discussion about this topic. Yet, we acknowledge that some participants may have avoided vulnerable issues regarding this unprecedented time of the pandemic. Finally, the interviews were held during the first part of a continuously evolving pandemic. Undoubtedly, this on-going crisis will continue to generate novel adaptations to learning and care that require investigation.

## Conclusion

For participants, working on the COVID-19 frontlines was hugely influential for both their learning and professional identity development. Our results reiterate the need to create learning environments where medical students are afforded ample opportunities to both participate in hands-on learning and feel like a valued member of the care team. Our participants told us that such opportunities do not have to be extraordinary or overly complex. That is, for medical students who primarily observe rather than play an active role in traditional clinical settings, even rudimentary [yet critically important] tasks were perceived as exciting and valuable for learning. We remind practicing physicians to remember how impactful simple clinical tasks and collaborative moments are for professional development— particularly for learning skills like communication that can be challenging to acquire and finesse without direct engagement.

## Data Availability

All data produced in the present study are available upon reasonable request to the authors

## Declaration of interest

The authors declare no conflicts of interest.

## Notes

### Competing Interest Statement

The authors have declared no competing interest.

### Funding Statement

This study did not receive any funding.

### Author Declarations

Ethics committee of EKNZ (Ethikkommission Nordwest- und Zentralschweiz) gave ethical approval for this work (Req-2021-00518).

