## Supplementary material for "Medical Students’ Perceptions of Learning and Working on the COVID-19 Frontlines: “… a confirmation that I am in the right place professionally.”": Figure 1

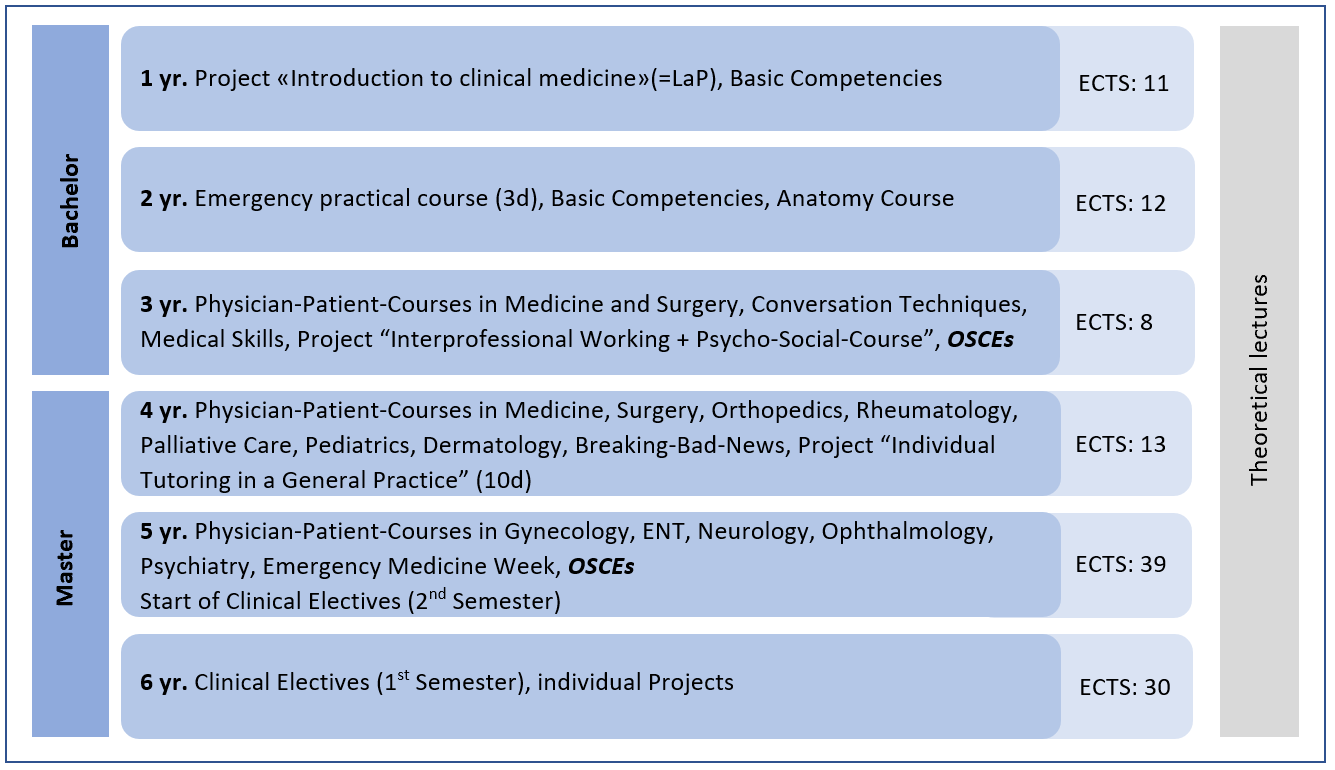

Figure 1: Clinical and Hands-on Courses at the University of Basel (Detailed description: Medical students gain their first clinical experiences through a format called Learning on the project (LaP) in the first year. LaP includes several bedside-teaching sessions and other learning opportunities in the hospital such as observing surgery. Before or during the first year, the University also expects a month-long clerkship as a nurse assistant in a hospital. In the second year, medical students work three days in the ED, guided by nurses. From the third year to the fifth year, Basel's medical students participate in the "physician-patient-courses" to learn conducting a medical history and physical exams. Those small group learning sessions cover some of the most critical fields in medicine, closely supervised. In the fourth year, medical students observe a family doctor to collect further professional insights. From the fifth to sixth year, medical students work as full-time clerks in the hospital for one year. As the final exams of the third and fifth year, *objective structured clinical examinations (*OSCEs) are conducted to monitor their clinical competence skills and development.)
